# Vaccination and collective action under social norms

**DOI:** 10.1101/2024.04.08.24305497

**Authors:** Bryce Morsky

## Abstract

Social dynamics are an integral part of the spread of disease affecting contact rates as well as the adoption of pharmaceutical and non-pharmaceutical interventions. When vaccines provide waning immunity, efficient and timely uptake of boosters is required to maintain protection and flatten the curve of infections. How then do social dynamics affect the timely uptake of vaccines and thereby the course of an epidemic? To explore this scenario, a behavioural-epidemiological is developed here. It features a tipping-point dynamic for the uptake of vaccines that combines the risk of infection, perceived morbidity risk of the vaccine, and social payoffs for deviating from the vaccination decision making of others. The social payoffs are derived from a social norm of conformity, and they create a collective action problem. A key finding driven by this dilemma is that waves of vaccine uptake and infections can occur due to inefficient and delayed uptake of boosters. This results in a nonlinear response of the infection load to the transmission rate: an intermediate transmission rate can result in greater prevalence of disease relative to more or less transmissible diseases. Further, global information about the prevalence of the disease and vaccine uptake increases the infection load and peak relative to information restricted to individuals’ contact networks. Thus, decisions driven by local information can mitigate the collective action problem across the population. Finally, the optimal public policy program to promote boosters is shown to be one that focuses on overcoming the social inertia to vaccinate at the start of an epidemic.

## 1 Introduction

Human behaviour modulated by social norms and other informal social rules is a major driver of disease spread [12, 22]. The uptake vaccines and the use of masks can be determined by social norms and group identity. Herd immunity suffers from a collective action problem, since non-vaccinators can free-ride on the vaccination of others. Because such social factors are so important to the spread of disease, it is critical to incorporate them into disease models. There is a wealth of literature on epidemiological models addressing this research topic [1, 10, 13, 17, 20, 21, 45–47, 49, 50]. And, during the COVID-19 pandemic, this literature has grown immensely. These models include individuals’ awareness of the the death rate or infection rates [32, 48] as well as individual decision making about uptake of non-pharmaceutical interventions [32, 37, 39]. Such models have shown that public policy makers face difficult decisions in light of the role of social dynamics. One example is that optimal social distancing levels can be highly sensitive to the *R*_0_ of the disease [32], and another is that misinformation can undermine the efficacy of public policy [43]. More generally, public policy makers cannot assume that individuals are purely rational decision makers. Rather, their decision making will be impacted by social and psychological factors. In the economics literature, these have been modelled by social payoffs (or relational utility), which have been shown to fundamentally change the qualitative nature of games and decision making [9, 44]. Social payoffs can measure feelings of guilt, joy, anger, and frustration that are generated by social interactions. With respect to the spread of disease, these could be generated by individuals’ decisions to be vaccinated or use non-pharmaceutical interventions given the behaviours of their friends, family, and others in society.

Heterogeneity, and in particular heterogeneous behaviour, is another fundamental determinant of disease spread. It is well-established that heterogeneity in contact patterns [5] and age structure [28] can dramatically affect infectious disease dynamics. Likewise, individual variation in susceptibility to disease can also alter how a disease spreads through a community and what the final distribution of outcomes are. Individual risk perception can also vary when it is determined by an individual’s social environment. In the example of ring vaccination, local dynamics can be beneficial in reducing the spread of disease: individuals with infectious neighbours choose to vaccinate to protect themselves and thereby overcome the social dilemma arising from a voluntary vaccination policy [11, 31, 34, 35].

An important epidemiological concern that can be influenced by social factors is waning vaccine immunity given an endemic disease [15, 16, 18]. How do social dynamics impact the uptake of booster shots and what recommendations are there to public policy makers to minimize the spread of disease? To explore these questions, this paper presents a compartmental epidemiological model that incorporates social behaviour, waning vaccines, and vaccine boosters. Individuals weigh the pros and cons of deciding to be vaccinated/-boosted while knowing the current infection rate of either their contact network on which the disease spreads or the population as a whole. The vaccine is assumed to include a real or perceived cost (this could be monetary or a risk of morbidity). Additionally, a negative social payoff (i.e. a social cost) is generated from deviating from others’ behaviour, since individuals are assumed to be conformists. It has been found that such conformity can result in non-monotonicity of the attack rate with respect to key epidemiological parameters such as the transmission rate the and the efficacy of non-pharmaceutical interventions [32]. Together, these factors determine individual’s vaccine decision making, and thus the course of the disease.

In addition to understanding how such dynamics impact the spread of disease, this paper explores the role of public policy in promoting boosters. Specifically, optimal control theory, which has been widely applied to epidemiology [2–4, 14, 19, 29, 42], is employed to find the optimal strategy in promoting the vaccine to the total amount of infections over time.

## 2 Methods

### 2.1 ODE model

Consider a compartmental epidemiological model coupled with a behavioural dynamic to model the rate of vaccine uptake by the population. The compartments for individuals include susceptibles, infectious, and protected. By assuming that natural and vaccine immunity are equivalent, the protected class represents both recovered individuals and those who have been vaccinated. The system of differential equations then takes the form:

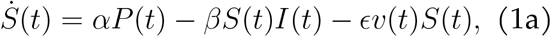

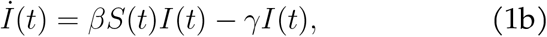

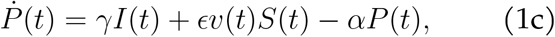

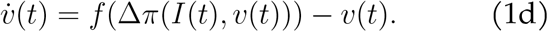

Here *S*(*t*), *I*(*t*), and *P* (*t*) are the frequencies of susceptibles, infectious, and protected at time *t*, respectively. *α, β*, and *γ* are the resusceptibility, transmission, and recovery rates, respectively.

The state variable *v*(*t*) ∈ [0, 1] is the degree to which individuals’ want to be vaccinated. 1*/*ϵ represents the time it takes for an individual to be vaccinated (e.g. the time until a vaccination appointment can be met), and thus ϵ*v*(*t*) is the vaccination rate. The change in *v*(*t*) is determined by the Granovetter-Schelling social dynamic, Equation 1d. This dynamic was originally developed in the social science literature to model collective action [23–26, 30, 40, 41], and has recently been employed to model the uptake of non-pharmaceutical interventions during an epidemic [32]. *f* (Δ*π*(*I*(*t*), *v*(*t*))) ∈ [0, 1] is a smoothed best response function of the difference between the payoffs to choosing to be vaccinated and choosing not be vaccinated, Δ*π*(*I*(*t*), *v*(*t*)) (which in turn is a function of the prevalence of infection and vaccination rate as discussed below). This best response function represents the probability that an individual will choose to be vaccinated, and thus is an increasing function with respect to Δ*π*, and is assumed to be a sigmoid function. An example of such a function and the one used in the numerical simulations in this paper is

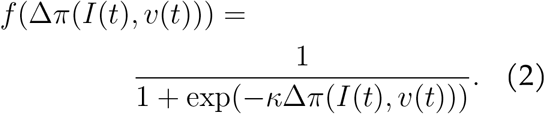

*κ* > 0 is the sensitivity to the payoff difference. The greater it is, the sharper the transition in vaccination decision-making.

When comparing the payoff difference between being vaccinated and not, Δ*π*(*I*(*t*), *v*(*t*)), individuals weigh the costs and benefits of choosing to be vaccinated including social ones. Specifically, individuals will consider their risk of being infected (frequency of infectious, the transmission rate, and whether or not they are vaccinated), the (perceived) cost of the vaccine (this can be monetary or a risk of morbidity), and a social cost for deviating from the behaviour of others. The latter is derived from assuming a social norm of conformity: individuals pay a social cost for not behaving like the average behaviour in the population. Social costs derived from norms in this way have been studied within the psychological game theory literature [6–9, 38, 44]. Summing together these components, the payoff for vaccinating 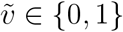 ∈ *{*0, 1*}* is

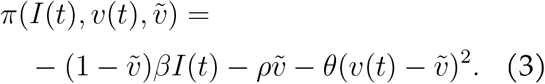

It is assumed that the protection provided by the vaccine wanes, but that there is no vaccine failure, hence the first term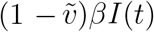). *ρ* is the risk or cost to be vaccinated. 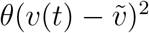 is the social cost, which is positive if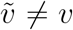 . The payoff difference is therefore

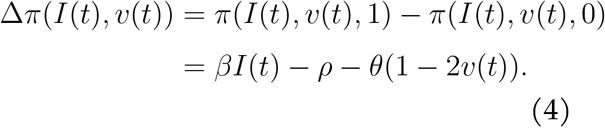

A similar type of payoff difference has been previously used to model the adoption of non-pharmaceutical interventions, which change the transmission rate [32].

### 2.2 Agent-based model

In addition to the above system of ODEs, an agent-based small-world network model is also considered here. The network, an Erdős-Rényi random network, plays two roles. For one, the disease can spread along it node to node. For another, it determines the information that individuals have and the social pressures they experience (individuals pay a social cost from deviating from the behaviour of their neighbours). Thus the payoff comparisons individuals make between choosing to be vaccinated or not depend on the infection status and preferences of an their neighbourhood, which is either their immediate neighbours (the local scenario) or all individuals in the network (the global scenario).

Each turn in the agent-based model is one day, and individuals are selected in a random order to determine the outcome for them that day. On day *t*, a susceptible individual *n* is infected with probability

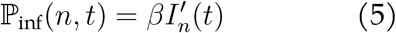

where 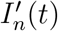 is the frequency of infectious individuals in *n*’s local neighbourhood on day *t* (i.e. those with whom they share an edge). If a susceptible is not infected, they may choose to be vaccinated/boosted. When making this decision, individuals observe the infection status and vaccination preferences of their neighbourhood, 𝒩 . Using Equation 2, individual *n* chooses to vaccinate with probability

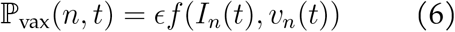

where *f* is Equation 2. *I*_*n*_(*t*) is the frequency of infectious individuals in *n*’s neighbourhood (e.g. 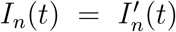 in the case of local information). *v*_*n*_(*t*) is the average preference to be vaccinated in *n*’s neighbourhood. Specifically,

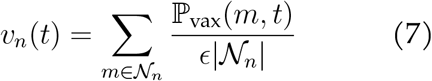

where 𝒩_*n*_ is *n*’s neighbourhood and |𝒩_*n*_| its size. Note that any infectious or protected individuals in *n*’s neighbourhood affect these values, since it is their opinions rather than their statuses that affect *n*’s preference. Each day an infected individual may recover with probability *γ*. And, a protected individual will lose protection and become susceptible with probability *α*.

For both simulations of the agent-based model and numerical solutions of the system of ODEs, the parameter values from Table 1 were used. *α, β*, and *γ* were chosen to roughly match the values for the SARS-CoV-2 virus. An exposed category for infected individuals is omitted for analytical tractability and because numerical simulations showed qualitatively similar results. The mean node degree for the agent-based model is *δ*, which was explored over the values 10 and 100 in a population of 10, 000 individuals. Though vaccines can be administered through walk-ins, the delay of 1*/*ϵ = 3 was chosen to represent a delay between when an individual decides to be vaccinated and when they may schedule and make an appointment. *κ* = 1000 is chosen so that there is a smooth but abrupt change in behaviour. A magnitude higher or lower *κ* has a marginal effect on the results. Finally, the magnitude of *θ* and *ρ* were taken from [32], which were found to be the range in which oscillations in preferences during an epidemic can occur.

**Table 1:** Parameters for numerical simulations.

| Parameter | Definition |
| --- | --- |
| $1/\alpha = 100$ days | resistance period |
| $\beta = 0.4/\text{day}$ | transmission rate |
| $1/\gamma = 7$ days | recovery period |
| $\delta$ neighbours | mean node degree |
| $1/\epsilon = 3$ days | time to be vaccinated |
| $\kappa = 1000$ | payoff sensitivity |
| $\theta = 0.01$ | relational cost |
| $\rho = 0.01$ | risk of vaccine morbidity |

## 3 Results

### 3.1 Equilibria analyses

First consider the ODE model. Given that *S*(*t*) + *I*(*t*) + *P* (*t*) = 1, we may reduce it to the three dimensional system:

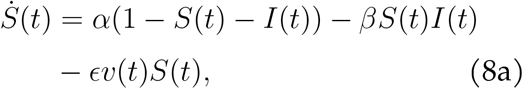

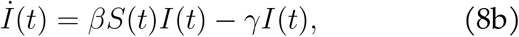

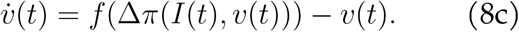

Here, we find the equilibria of this reduced system and analyze their stability. To start, consider the disease free equilibrium (DFE). There are a set of DFE corresponding to different equilibrium vaccination rates: 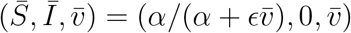 .

#### Theorem 3.1

*If* 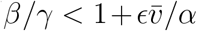 *and ∂f*/*∂v* < 1 *at a DFE, then it is stable*.

*Proof.* Let *f* = *f* (Δ*π*(*I*(*t*), *v*(*t*))) for notational simplicity. Linearizing about the DFE, we have the following Jacobian matrix:

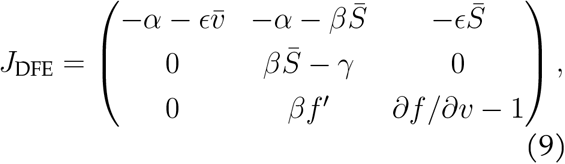

which has eigenvalues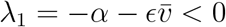,

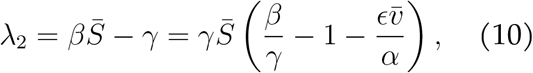

and *λ*_3_ = *∂ f/∂ v* − 1. Thus the DFE can be stable so long as: 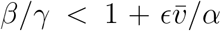 and *∂ f/∂ v* = 2*θf*^*′*^ *<* 1. Since *f ∈* [0, 1] is a sigmoidal function, there may be at most three equilibria for 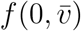 for 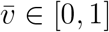 and a minimum of one. And for at least one of these equilibria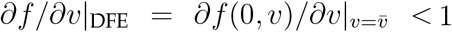.

Next consider the endemic equilibrium (EE). Here, 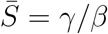 and

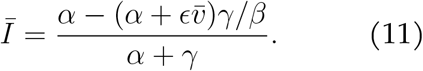

Note that *Ī* ∈ (0, 1) if and only if 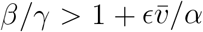, which leads to the next theorem.

#### Theorem 3.2

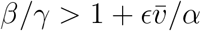, *∂f/∂v <* 1, *and* 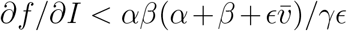 *at an EE, then it is stable*.

*Proof.* Linearizing about the EE, we have the following Jacobian matrix

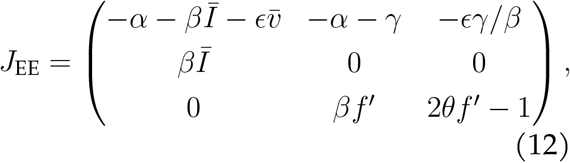

and characteristic polynomial

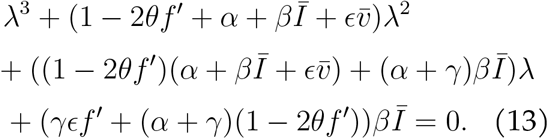

Note that if *∂f/∂v* = 2*θf*^*′*^ *<* 1, then all of the coefficients are positive, which is a necessary though not sufficient condition for stability by the Routh-Hurwitz criteria. We further require that *c*_2_*c*_1_ − *c*_0_ *>* 0 for coefficients *c*_*i*_ of *λ*^*i*^ as follows:

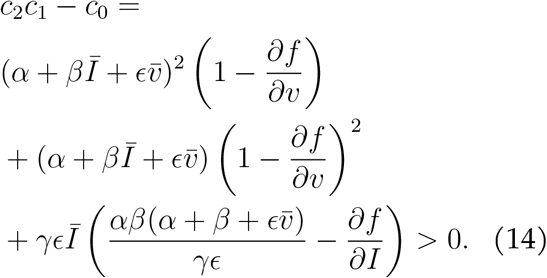

Conversely, *∂f/∂v* > 1 or 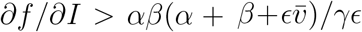 are necessary conditions for an EE to be unstable.

When there are no stable DFE and EE, cycles can occur. Figure 1 depicts an example.

**Figure 1.**
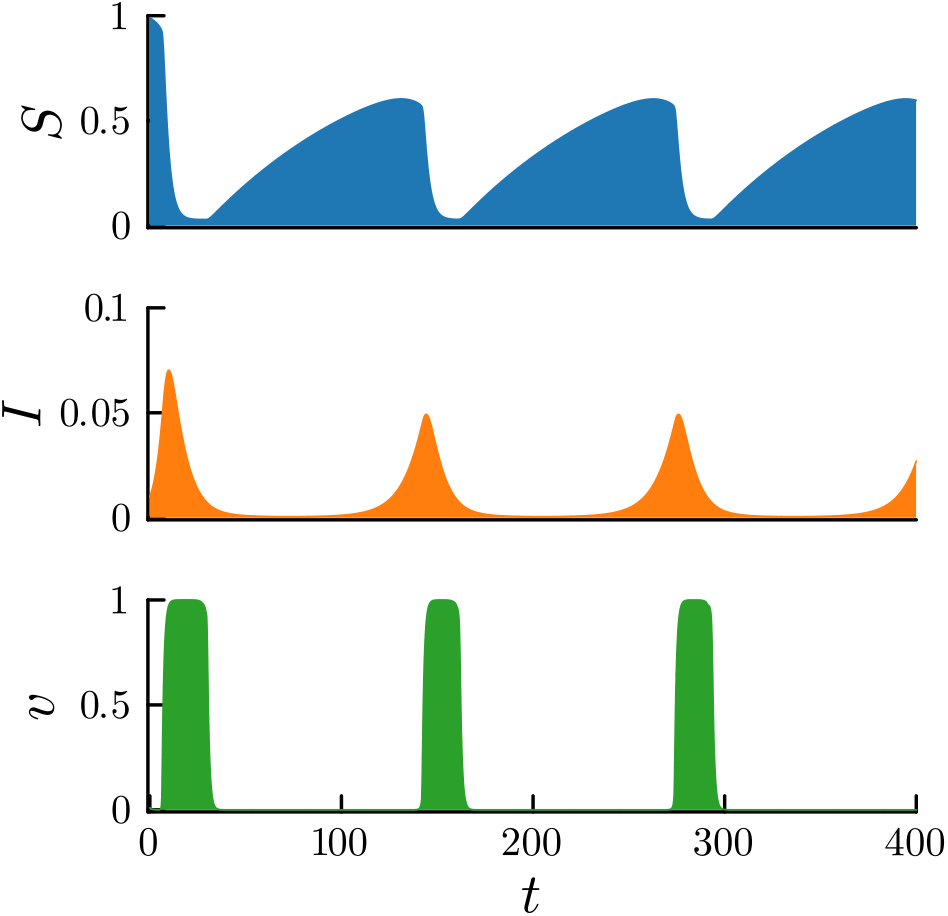
A representative example of observed cycles. Rising infections sway susceptibles to become vaccinated, which drives down infections. In turn, the vaccination rate declines. As immunity wanes, susceptibles rise and the process repeats. Initial conditions are *S*_0_ = 0.99, *I*_0_ = 0.01, and *v*_0_ = 0.01 and parameter values are taken from Table 1.

With an initially highly susceptible population, the infection spreads inducing vaccination. However, due to the social cost of not conforming, vaccination rates rise more slowly than they would only computing the risk of the infection and the cost to vaccinate. Once vaccination rates are high, they remain so even after the disease has been controlled. In this case, conformity aides in sustaining vaccination rates. Once the vaccination rates do drop, susceptibility rises due to waning immunity, which leads to another outbreak.

### 3.2 Parameter analyses

Figure 2 shows the effects of various parameters on the infection load, i.e. the total amount of infectious individuals. The infection load is calculated by summing up the number of infectious from time *t* = 0 to *t* = 400 in increments of 1. In each panel, a single parameter is varied while the remainder are taken from Table 1.

**Figure 2.**
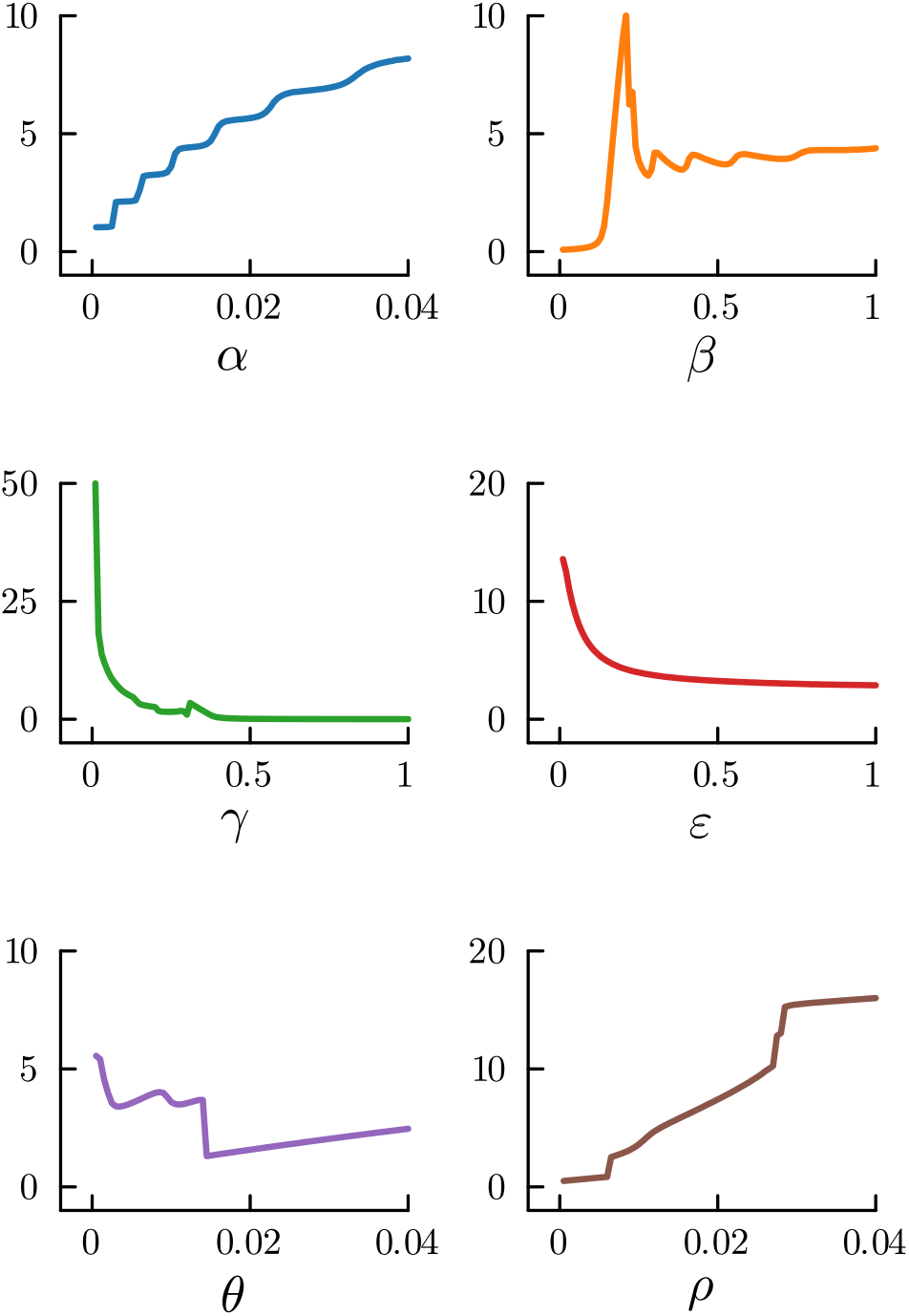
Infection load (*y* axes) for varying parameters *α, β, γ, ϵ, θ*, and *ρ*. When varying a parameter, the others are fixed and taken from Table 1.

Recent studies have shown nonmonotonicity in outcomes from epidemics for changing parameters [32, 37], which is also observed for several parameters here. For example, the highest infection load occurs for an intermediate transmission rate *β*. When *β* is sufficiently low, individuals do not want to be vaccinated. Thus, increasing *β* will increase the infection load up until the vaccine is widely desired at which point the infection load drops. In addition to this general observation, there are also smaller variations in the infection load for varying *β*; specifically, a sawtooth like pattern. This result is due to the tipping point dynamic, which leads to the nonlinear uptake of vaccines and thus the non-monotonicity observed.

Varying the social cost *θ* also results in non-monotonicity, since there is a trade-off between low and high values of it. If *θ* is low, there is little social resistance to increasing the vaccine uptake when vaccinations are infrequent and infections are low but rising. At the start of an epidemic with a largely unvaccinated population, this is beneficial. On the other hand, there is also little social resistance to decreasing the vaccine uptake when infections are reduced, which reduces the rate at which individuals receive boosters and leaves the population susceptible to a resurgence of the disease. When *θ* is large, social resistance retards vaccine uptake at the start of the epidemic, but can sustain individuals receiving boosters. Altering the other parameters generally results in monotonic changes in infection load. Increasing *α* and *ρ* increases the infection load nonlinearly while increasing *γ* and *ϵ* decreases it.

### 3.3 Optimal control

Conformity due to the social norm results in a slow sub-optimal uptake of vaccines at the beginning of the epidemic. This is because the descriptive norm (the norm that describes the behaviour of others) is, correctly, that vaccine uptake is low. Can a public policy campaign that adjusts this belief alter the outcome of the epidemic when it is endemic? To explore this possibility, consider a control variable *u*(*t*) for the system that represents the degree to which public policy promotes individuals’ desires to be vaccinated. Assuming that there is some upper bound its strength, we have that 0 ≤ *u*(*t*) ≤ *u*_max_ ≤ 1. Payoffs are thus adjusted by replacing *v*(*t*) (the true vaccine preference of individuals) with (1−*u*(*t*))*v*(*t*)+*u*(*t*) (the promoted one) into the payoffs. The state equations for the optimal control problem are thus those of Equations 8a-8c with Δ*π*(*I*(*t*), *v*(*t*)) replaced with

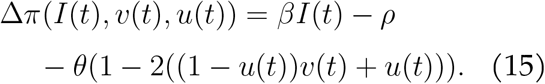

Assume then that we wish to minimize the infection load as well as the control *u*(*t*). This gives us the cost functional

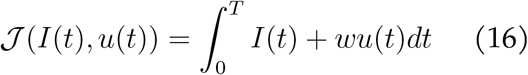

to minimize where *w >* 0 is the weighting of the control. Though the epidemic is endemic and thus the time horizon is infinite (*T* → ∞), we can truncate the time for the numerical solutions to *T* = 400. Using Pontryagin’s minimizing principle, we have the Hamiltonian

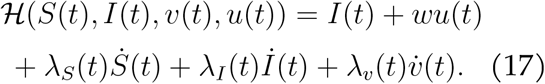

The optimality condition is

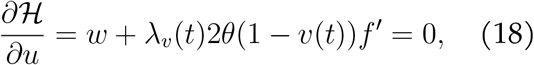

and the adjoint equations are

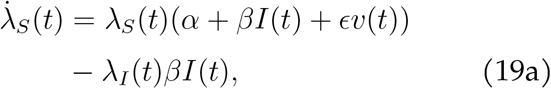

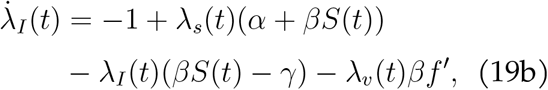

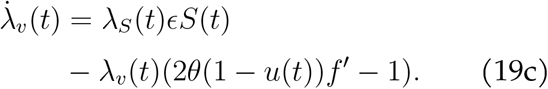

Since we are assuming a finite time horizon problem, we have the transversality conditions: *λ*_*S*_(*T* ) = *λ*_*I*_(*T* ) = *λ*_*v*_(*T* ) = 0.

Figure 3 depicts a numerically solution using Julia and the InfiniteOpt.jl package [36] for the optimal control along with the resulting trajectories for the state variables for the first 200 days. The weight and maximum control are *w* = 0.01, and *u*_max_ = 0.8, respectively. All other parameters are taken from Table 1. At the start of the epidemic, the control is maximized to overcome social resistance to vaccination and thereby blunt the initial spread of the disease. Once vaccination rates are high, the control can be emoved, since vaccinations are bolstered by social pressure. As this pressure wanes, the control must be reintroduced at several times as the vaccination levels dip to sustain boosting and control of the disease. Long term, this must be sustained to prevent reemergence of the disease.

**Figure 3.**
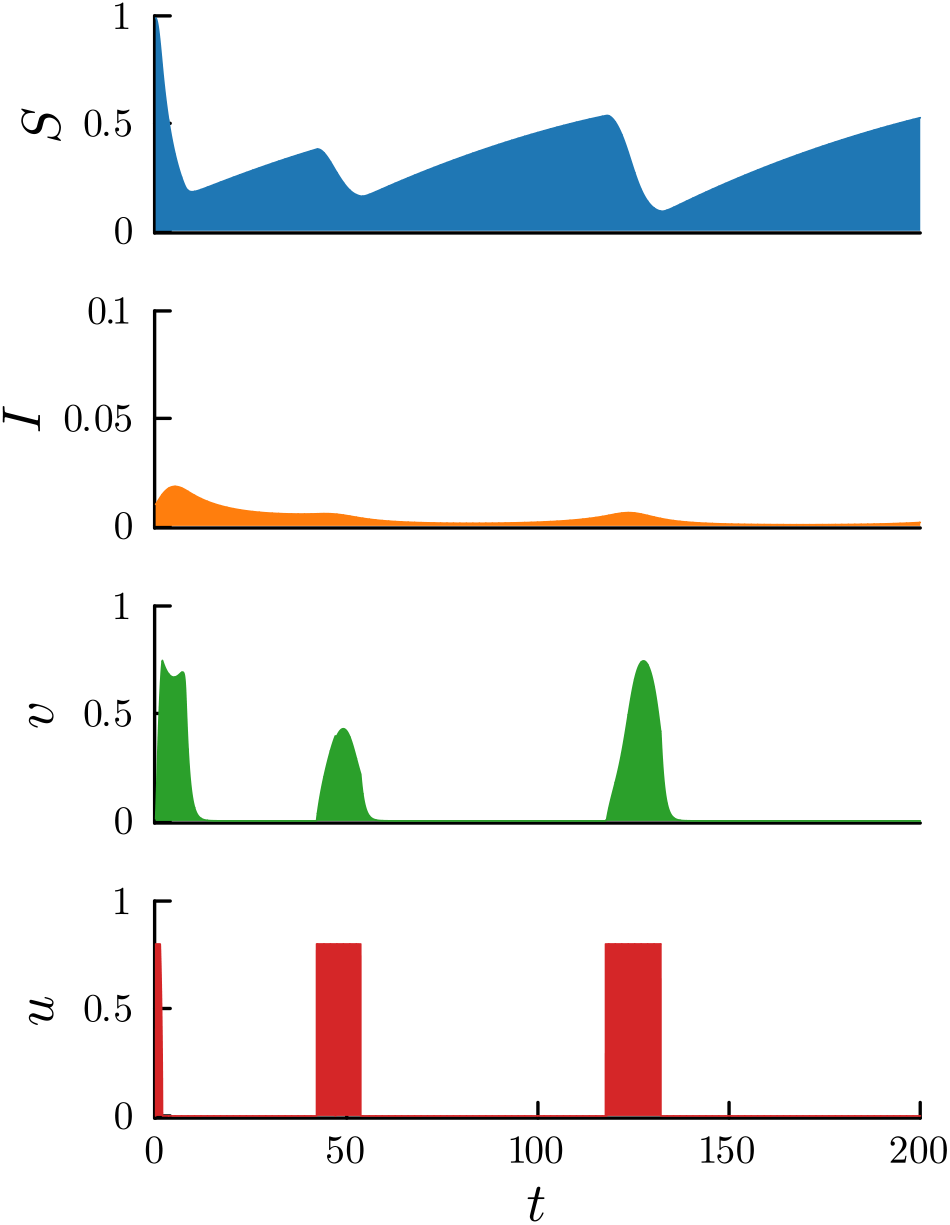
Numerically solved optimal control solution. Initial conditions are *S*_0_ = 0.99, *I*_0_ = 0.01, and *v*_0_ = 0.01. *w* = 0.01 and *u*_max_ = 0.8. Other parameter values are taken from Table 1.

### 3.4 Networks and heterogeneity

Here is reported the results for the network model that incorporates heterogeneity. Figure 4 depicts representative time series for the different cases of local or global knowledge of the disease and vaccine uptake rates as well as different neighborhood sizes (withe mean node degree *δ* = 10 and 100). The population size is 10, 000 individuals with initially 1% infected and the preference to be vaccinated at 0.01 for all individuals.

**Figure 4.**
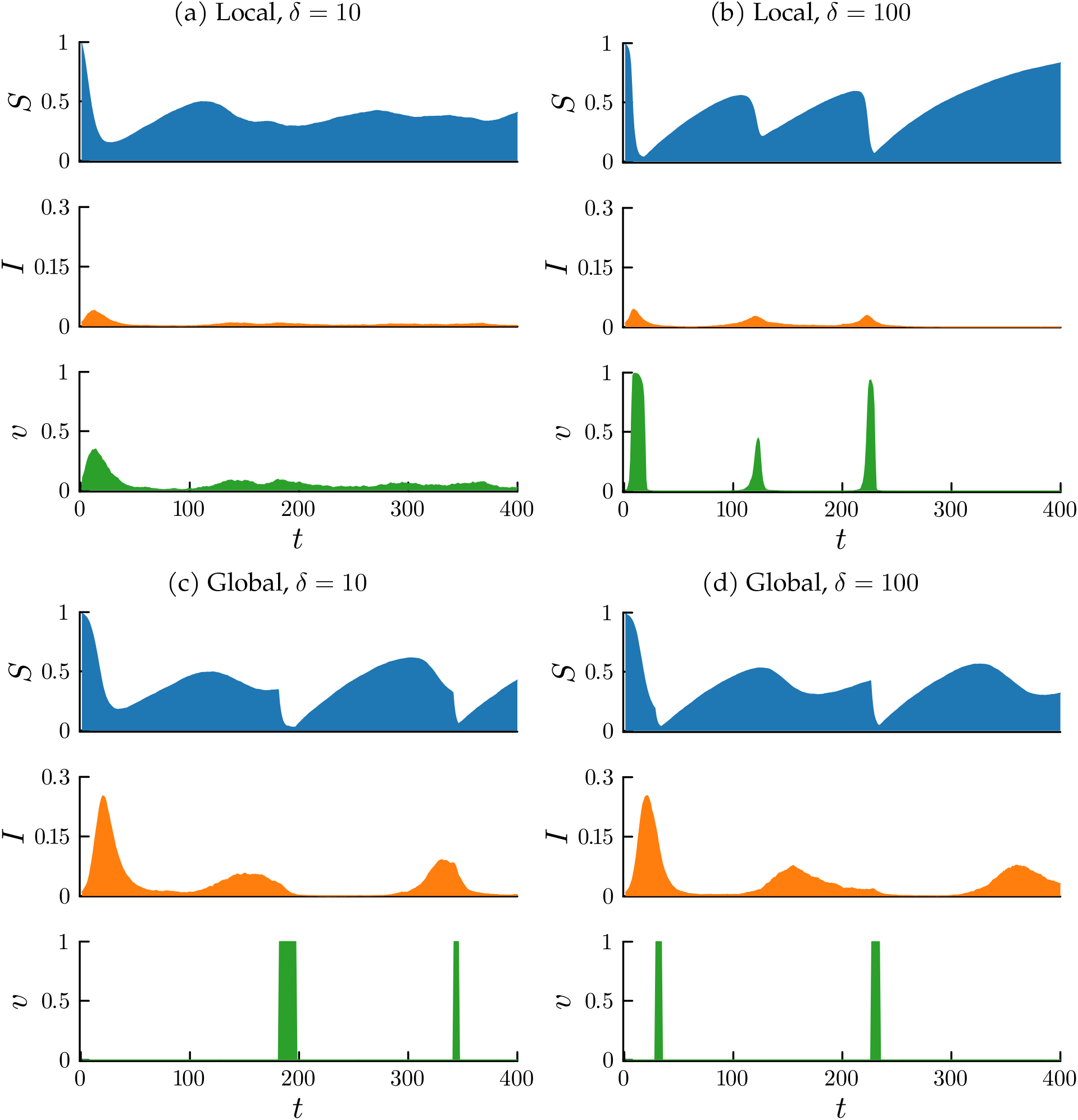
Representative time series for the network model with local or global knowledge of disease prevalence and vaccine uptake levels. Initial conditions are *S*_0_ = 0.99, *I*_0_ = 0.01, and *v*_0_ = 0.01. Parameter values are from Table 1.

Localness results in fewer infections and flatter waves of infection than globalness. For individuals near the locus of the infection, the perceived and real risk of being infected is high and thus they vaccinate preventing the spread of disease to others. This phenomenon is known as ring vaccination and is a means by which voluntary vaccination strategies can be effective [27, 31, 33–35]. More information leads to steeper switches in desires to become vaccinated as can be seen in panels b-d. Individuals become coordinated and synchronized in their decision about whether or not to be vaccinated, which results in larger swings in vaccine uptake levels and peaks in infections. The social payoff from the norm retards uptake of the vaccine, initially. This effect allows the disease to progress more than under local information, which spreads out the decision making and vaccination in space and time preventing these swings of behaviour.

## 4 Discussion

This paper has explored the effect of a social norm of conformity on decisions to vaccinate when the vaccine has a waning im-munity. It has been shown that this can lead to both disease free and endemic equilibria as well as a nonlinear response to varying parameters. Further, it has shown that there is an optimal public policy in promoting the vaccine, which begins with a strong promotion to overcome the “norm stickiness” [32] followed by a relaxation and then a return to promote boosting at regular intervals to keep the disease under control.

Mirror results in [32, 37], varying key parameters of this system is non-monotonic and nonlinear. In particular, changing the transmission rate does not always increase the number of infectious individuals. For a high transmission rate, individuals will regularly receive boosters thus limiting the spread of the disease. However, there is an intermediate level of transmission in which the social dynamic frustrates efficient and timely uptake of the vaccine resulting in a high infection load. Another key parameter with a nonlinear response to variations is the weight of the social cost *θ*. For a very low social cost, there is little social impediment to being an early adopter of the vaccine. However, there is also little social pressure to sustain adoption of boosters. Increasing the social cost from such a low level can be beneficial. Since, although it impedes the initial adoption of the disease, it promotes sustained boosters and thus reduces future waves of infection.

Heterogeneity in behaviour stemming from heterogeneity in information and local conditions can promote a low infection load and flatten the curve of infections. Global information, on the other hand, synchronizes the behaviour of individuals leading to mass uptake of the vaccine as well as mass abandonment of receiving boosters. This effect results in a greater infection load and larger peaks of infections as vaccination levels wane and individuals choose to be vaccinated too late in response to a resurging epidemic. Boosters are thus taken in response to rising infections rather than in preparation for them. Nonetheless, global information may be useful in some scenarios not covered here. Synchronization of behaviour may be effective when information is inaccurate or when the disease can be eradicated. Though shaping of information has been explore to some degree here, future work could more deeply explore this idea.

A further limitation of the model and direction of future research include the assumption that vaccination decision making is not modulated by the duration of the epidemic. The population can repeatedly be activated to choose to be vaccinated. In reality, individuals could become disillusioned with receiving boosters regardless of the levels of infections in the population. This could be especially true if there is inaccurate information and group dynamics that drive vaccination decision making. Here individuals are “rational” with respect to social pressures, and these pressures are essentially the same. Although, in the agent-based model, they can vary as the neighbourhoods vary. However, individuals could have different social pressures depending on different social norms. e.g. individuals may only wish to conform with those who share a similar identity to them. Or, they may be driven to take the opposite behaviour of those different from them. There are many such scenarios that could be explored.

Given the model and results here, what recommendations are there for public pol-icy makers? For one, when vaccine uptake is low and uncommon, vaccination should be promoted heavily and to a degree greater than the current infection rates warrant so as to overcome social resistance. However, resources used for such promotion can be saved once vaccination becomes common, since social dynamics can sustain it in the short term. As the disease prevalence decreases, however, it is key to promote the vaccine again to prevent following waves of infection. Additionally, it may be useful to promote vaccination more locally than globally. Though this may be a futile effort, if individuals’ behaviours can be decoupled from the global dynamics and the behaviours of peers outside of their locality, that would promote a steadier and more localized uptake of the vaccine and promote fewer infections.

## Data Availability

There is no data to report.

